# Modeling the dynamics of COVID19 spread during and after social distancing: interpreting prolonged infection plateaus

**DOI:** 10.1101/2020.06.13.20130625

**Authors:** Natalia L. Komarova, Dominik Wodarz

**Affiliations:** Department of Mathematics, University of California Irvine, Irvine, CA 92697; Department of Population Health and Disease Prevention, Program in Public Health, Susan and Henry Samueli College of Health Science, University of California Irvine, Irvine, CA 92697

## Abstract

Non-pharmaceutical intervention measures, such as social distancing, have so far been the only means to slow the spread of COVID19. In the United States, strict social distancing has resulted in different types infection dynamics. In some states, such as New York, extensive infection spread was followed by a pronounced decline of infection levels. In other states, such as California, less infection spread occurred before strict social distancing, and a different pattern was observed. Instead of a pronounced infection decline, a long-lasting plateau is evident, characterized by similar daily new infection levels. While these plateau dynamics cannot be readily reproduced with standard SIR infection models, we show that network models, in which individuals and their social contacts are explicitly tracked, can reproduce the plateau if network connections are cut due to social distancing measures. The reason is that in networks characterized by a 2D spatial structure, infection tends to spread quadratically with time, but as edges are randomly removed, the infection spreads along nearly one-dimensional infection “corridors”, resulting in plateau dynamics. Interestingly, the plateau dynamics are predicted to eventually transition into an infection decline phase without any further increase in social distancing measures. Additionally, the models suggest that a potential second wave becomes significantly less pronounced if social distancing is only relaxed once the dynamics have transitioned to the decline phase. The network models analyzed here allow us to interpret and reconcile different infection dynamics during social distancing observed in various US states.

## Introduction

The COVID19 pandemic has caused significant mortality and morbidity around the world [1], and the only means to slow its spread has been the implementation of non-pharmaceutical intervention methods, most notably social distancing [2, 3]. In the United States, stay-home orders have been given and have been implemented to various extents in the different states, which has resulted in an overall reduction of disease burden [4]. Economic considerations, as well as social distancing fatigue in the population, however, lead to the relaxation of non-pharmaceutical interventions. This in turn raises questions about the potential severity of a second wave of infection spread.

As these events are unfolding, it is useful to understand the dynamics of infection spread during social distancing, and how this might relate to a possible resurgence as the social distancing measures are relaxed. Mathematical models have been useful for understanding different aspects of COVID19 spread dynamics, e.g. [5-7], including the analysis of optimal strategies to relax social distancing measures [8]. Much of this work, however, is based on SIR models that are expressed in terms of ordinary differential equations [9], which assume that all individuals in a population mix with each other. When social distancing measures are in place, however, this assumption is likely violated. On a phenomenological level, it is possible to approximate the effect of social distancing by assuming a reduced transmission rate of the virus, because this is the net effect. On the other hand, to account for social distancing, it might be more realistic to consider modeling approaches that explicitly assume social connections among individuals, and that individuals are only connected to a subset of local contacts during stay-home orders.

Epidemiological data of COVID19 spread while social distancing measures are in place are characterized by some patterns that are difficult to explain by SIR models based on ordinary differential equations. In these models, exponential growth of the infection is observed until a peak is reached, after which a phase of exponential decline occurs. The observed COVID19 spread dynamics, however, have been found to be more complex. While exponential growth has been observed in certain locations, the infection spread seems to be better described by power laws in many other locations [10, 11], which could be brought about by the prevalence of local rather than more long-range interactions. In addition, when stricter stay-home orders are in place and the rate of infection spread is slowed down, different dynamics can be observed. In some states / counties, a pronounced decline of daily COVID19 cases is observed following strict social distancing. This tends to be the case if infection spread has been more severe, such as in New York (Figure 1a). In other locations, a relatively long-lasting plateau phase is observed, during which the number of daily cases seems to fluctuate around a steady average level (corresponding to a linear cumulative number of cases over time). This has been seen in places that implemented social distancing measures relatively early and controlled the spread effectively, such as in California or Washington State (Figure 1b). The standard SIR models that are based on ordinary differential equations cannot easily account for such a prolonged plateau.

**Figure 1:**
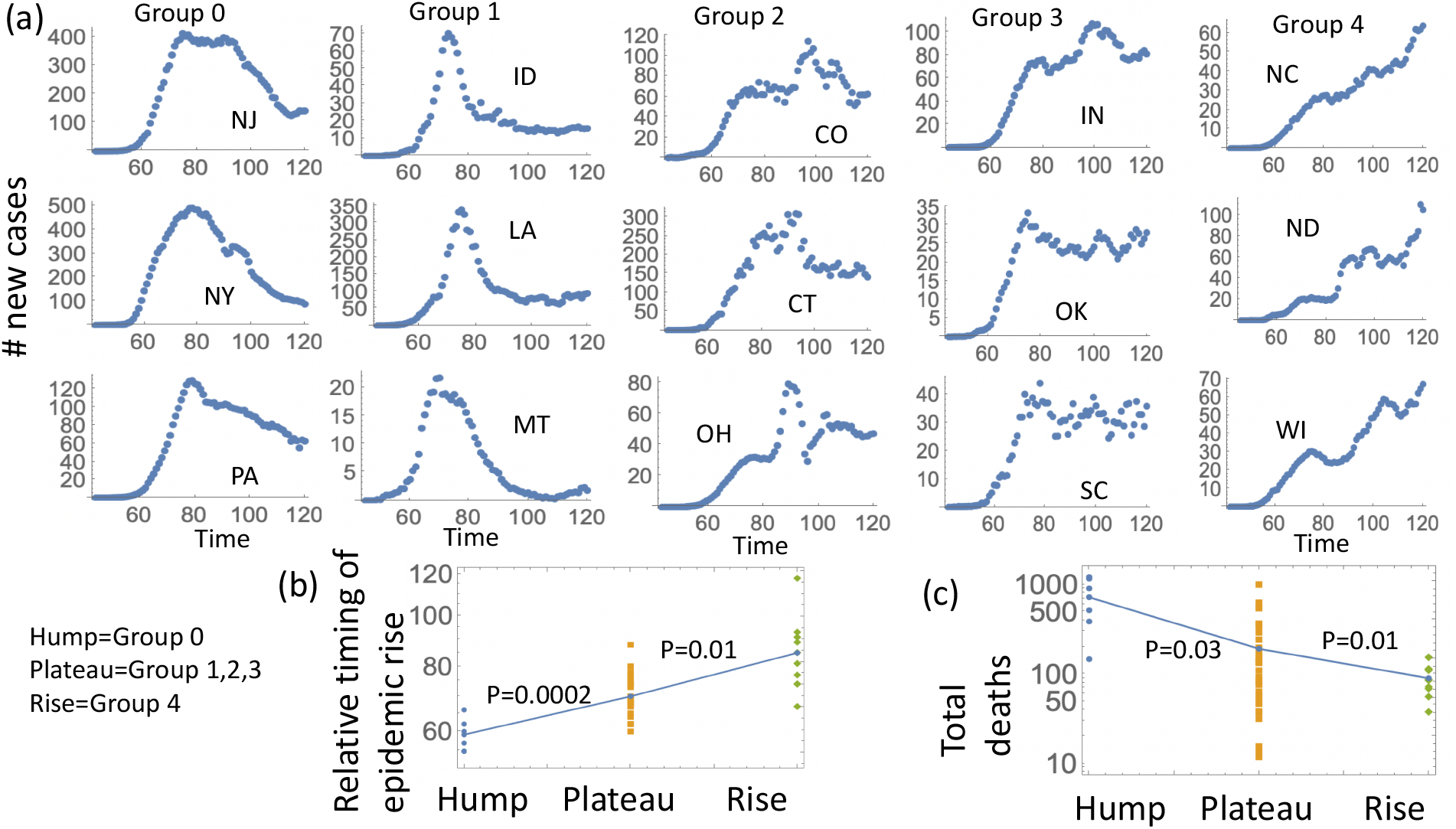
(a) Different patterns of COVID19 spread during social distancing across different states in the USA. Group 0 states show a relatively sharp decline of infections. Group 1 states show an initial decline, followed by convergence to a plateau. Group 2 states show a plateau without a significant decline during social distancing. Group 3 states show a rise, followed by a plateau. Group 4 states show a rise without convergence to a plateau. See Supplementary Materials for grouping methodology. (b) Correlation between the COVID19 spread pattern during social distancing with the relative timing of the epidemic rise (see Supplement Section 3 for details). A later rise of the epidemic is associated with a relatively early implementation of social distancing, which happens before the infection has spread significantly through the population. Thus earlier initiation of distancing correlates with the occurrence of a plateau or even a “rise” (which is thought to correspond to pre-plateau dynamics). Initiation of distancing after significant virus spread tends to correlate with a “hump”-shaped epidemic: a significant infection spread followed by a decline and lack of a plateau. (c) The same trend is seen when considering deaths as an indicator of the severity of infection when distancing is initiated. Less death correlates with the appearance of a plateau or a rise. More death correlates with a sharp rise of infection followed by a decline in the absence of a plateau.

Here, we analyze the spread of COVID19 using network models, which assume that individuals in a population do not all mix with each other, but that individuals interact according to contact networks. We assume the existence of contact networks both before and during strict social distancing efforts. Strict social distancing is implemented by cutting these network connections to varying degrees. We find that such models can reproduce the above-described intricacies of COVID19 spread. Hence, such models can give rise to the observed power law growth of infection spread, as well as the long-lasting infection plateaus observed during strict social distancing. In particular, if strict social distancing is put in place relatively early, the models predict a prolonged plateau phase during which the daily number of infections remain relatively constant. Interestingly, these dynamics transition naturally into a decline phase without any additional cutting of network connections (i.e. without stronger social distancing). In contrast, if strict social distancing measures are implemented only after the infection has spread to higher levels in the model, the plateau phase is less pronounced or absent, and a decline phase is observed right away. Consistent with previous modeling approaches [8], we find that the predicted second wave can be lower if social distancing is relaxed later. In contrast to the predictions from standard SIR models, however, our network models suggest that a lower second wave is only observed if social distancing is relaxed once the steady plateau phase is over and the number of daily new infections has started to decline. Several other differences are observed compared to standard SIR models when predicting how different parameters affect the magnitude of the second wave after social distancing measures are relaxed.

### A basic SIR model if COVID19 transmission

SIR models based on ordinary differential equations are a cornerstone of epidemiological infection models [9, 12], and they have also been an important component for COVID19 modeling [13]. They distinguish between susceptible individuals, S, infected, I, and recovered, R, individuals that are immune to infection. Also including a population of dead individuals, Z, the model can be written as follows:

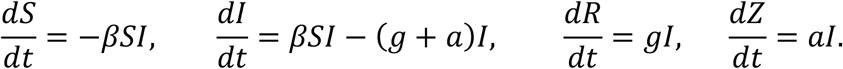

Reproduction or immigration of the host population is not included. Susceptible individuals are infected with a rate *β* upon contact with an infected person. Infected individuals die with a rate *a*, or recover with a rate *g*. The model assumes that recovered individuals cannot be infected anymore. There are no data yet indicating whether humans recovered from COVID19 are immune to secondary infections or not, although non-human primates have been shown to be protected approximately 30 days after onset of symptoms [14].

Because the model is given by ordinary differential equations, it assumes that all individuals in a population mix perfectly with each other. While this might not be realistic, social distancing can still be described by this model through a reduction in the infection transmission parameter β. Such a model has been investigated in detail in [8], but an analysis of this model is described in the Supplement, Section 1, to put our network models into context. In particular, according to the ODE model, delaying the start of social distancing leads to a lower infection level while distancing is taking place, but it may correspond to a lower final epidemic size after the relaxation of social distancing. Further, a longer duration of social distancing leads to a lower final epidemic size. Finally, stricter social distancing (lower infectivity, β) is predicted to result in a higher second wave infection peak and a higher final epidemic size. Similar results are also reported in [8].

## Network models

The same assumptions can be expressed in terms of network models [15-28]. These do not assume perfect mixing of individuals, but instead assume the existence of a certain number of social contacts per person, through which transmission can occur.

While the true structure of the human contact networks in the absence and presence of the pandemic are not fully understood, we analyzed two basic types of networks at different ends of the spectrum. In what we call a “spatial random network”, individuals are connected to their local neighbors. This might approximate a society under various social distancing measures, when people do not travel much. The degree of social distancing can be expressed by the average number of connections per individual. A network where individuals have a relatively large number of connections would correspond to a society that stopped traveling but is still interacting to a strong degree on a local level. Stricter social distancing measures would correspond to a reduced number of local connections in this network, e.g. due to people staying at home more. This network structure is shown schematically in Figure 2A. At the other end of the spectrum, we consider the scale-free Barabasi-Albert network [29], which is characterized by individuals having non-local connections, and a few individuals having a disproportionately large number of connections (Figure 2B). During social distancing, the connections in such a network can be reduced. Between these two scenarios, we consider a third network that we call “hybrid network” (Figure 2C), or a spatial scale-free network. The backbone consists of spatial network connections, with a set of long-range connections superimposed. Details of how these networks were constructed are given in the Supplementary Materials.

**Figure 2:**
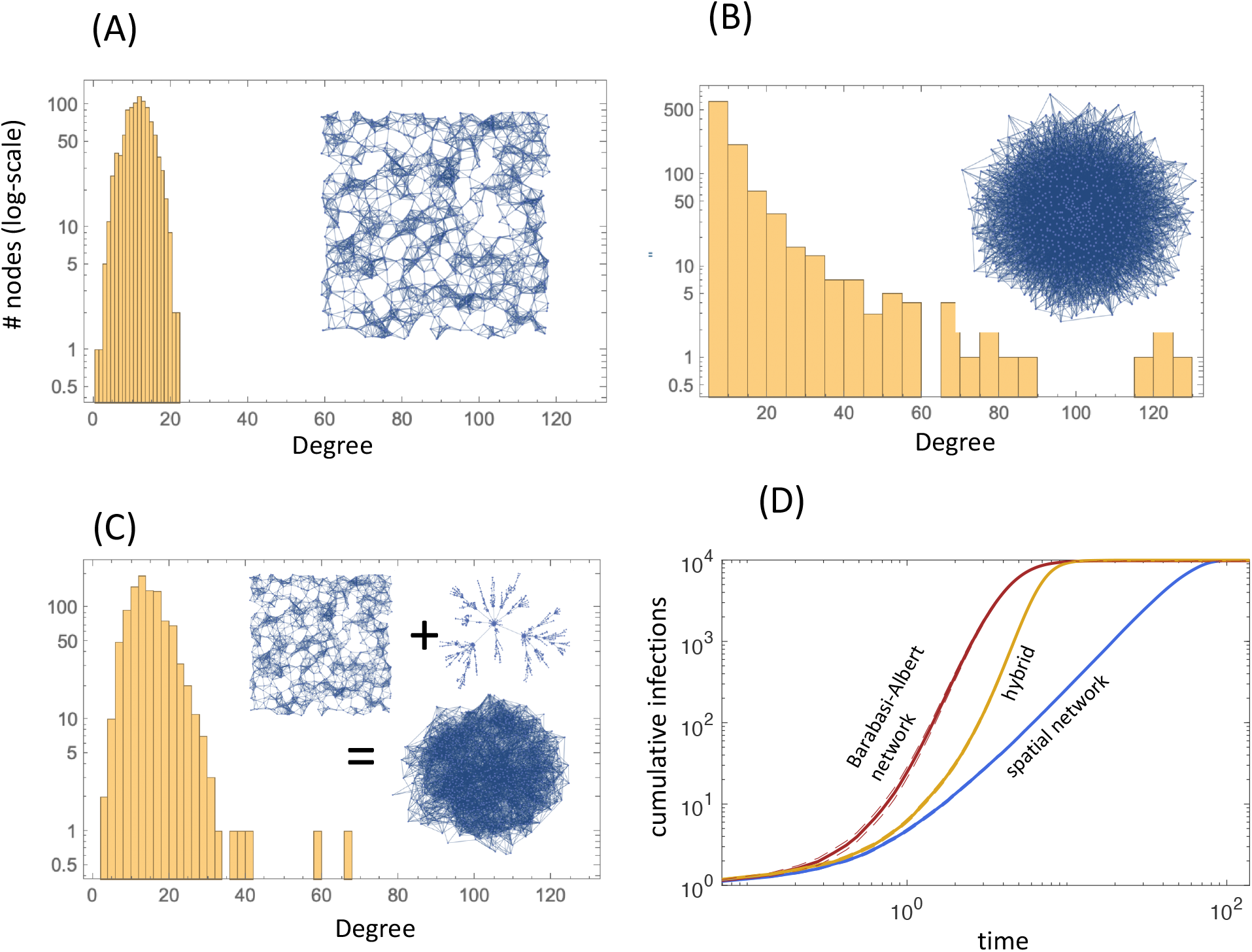
Different network types considered in this paper and their properties (See section 2.1 of the Supplement for details of construction). In (A-C), a typical degree histogram and a graphical representation of a typical network are presented. (A) A random spatial network, where nodes are connected largely to their neighbors, i.e. connections are short-range. (B) Scale-free Barabasi-Albert network, where no spatial correlations are found and there is a power law like tail in the degree distribution. (C) A hybrid network, in which a scale-free component is superimposed onto a spatial component. (D) Growth curves showing the infection spread in the three different networks. Standard errors are shown as dashed lines, which in some cases are too small to see. Parameters were chosen as follows. P_inf_=0.0001min^-1^ per edge; P_rec_=0.0001min^-1^; P_death_=0.00005min^-1^.

For each network type, we start with a given “null network”, which represents the state of society before strict local distancing measures. Stricter social distancing is implemented by randomly cutting network connections by a given percentage, which we can vary. Through this variation, we can consider a range of different intensities with which non-pharmaceutical interventions are implemented.

The infection dynamics on these networks were simulated stochastically with the following algorithm. Every time step, the network was sampled randomly until infected agents were selected M times, where M is the total number of currently infected individuals. For each infected individual that was selected, a death event occurred with probability P_death_, and a recovery event occurred with a probability P_rec_ (we refer to the probability of death or recovery as the probability of removal, P_removal_= P_death_ + P_rec_). With a probability P_inf_ x (number of social connections), an infection event was attempted. In this case, one of the connected individuals was chosen randomly for an infection event. If this individual was susceptible, an infection event proceeded. If this individual was either recovered or dead, no infection occurred. We note that in the context of the model, recovered and dead individuals have the same effect: they represent network nodes that are not available for transmission anymore. We will refer to these individuals as “removed” from the infection process.

### Basic growth laws

Here, we summarize the infection spread laws observed in the different network models assuming that the networks are in their “null state”, i.e. before connections are cut. The spatial model displays clear power law growth of the infection over time, which is due to the local connections that characterize this network (Figure 2D). The scale free Barabasi-Albert network displays an initial phase of exponential growth, followed by a transition to a power law, before the final epidemic size has been reached (Figure 2D). This behavior has been analyzed in detail before [20]. The hybrid network displays a similar behavior, although the growth is more skewed towards power law behavior (Figure 2D). Power law growth is not predicted by SIR models that are based on ordinary differential equations, and the observation of power law like spread of COVID19 across different locations [10, 11] thus indicates that network models might be more appropriate descriptions in many settings.

### Infection dynamics during social distancing

The network models considered here might shed light onto the mechanism underlying the observed prolonged plateau phenomenon discussed above (Figure 1). We first consider the spatial model. The simulation is started with the “uncut” version of the model that contains 10,000 agents that are characterized by a relatively large number of connections. When the number of infected individuals has reached 100 (1% of the total population), the simulation switches to a strongly cut version of the network, characterized by significantly fewer connections per agent. Figure 3A shows the dynamics averaged over many realizations of the simulation. Implementation of social distancing is followed by a peak of infections, after which the infection levels decline slightly and converge to a long-term plateau, during which average infection levels remain relatively constant. This is also reflected in a linear growth of the cumulative case counts in the simulation (Figure 3B). After a certain period of time at this plateau, the dynamics start to visibly decline. We note that this transition to the decline phase occurs without any further cutting of network connections, i.e. without any further implementation of non-pharmaceutical intervention measures.

**Figure 3.**
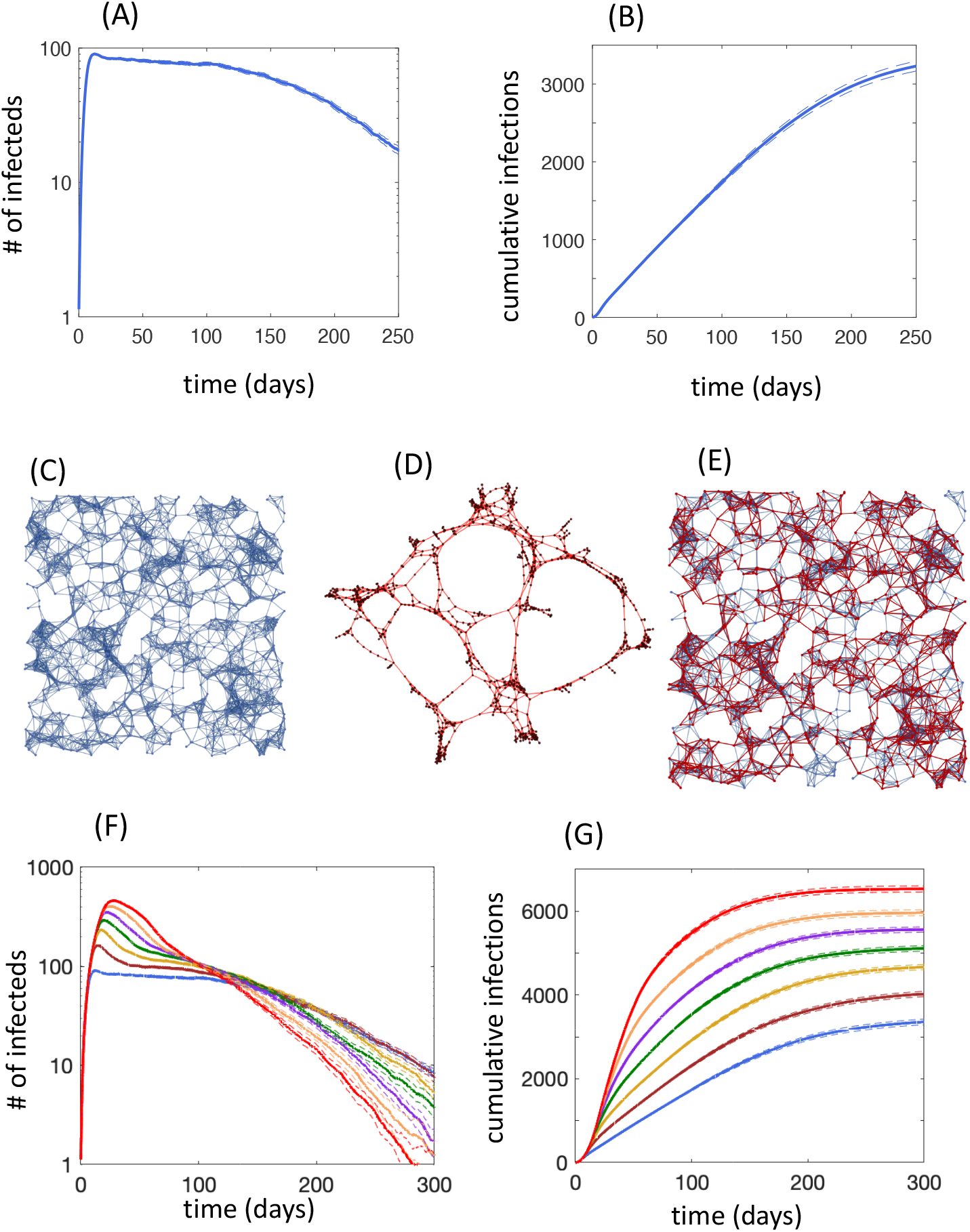
Social distancing dynamics in the spatial network. (A) Growth in the uncut spatial network occurs until 100 infected individuals are present, at which point half of the network connections are randomly removed. The average trajectory over 900 runs is plotted, and standard errors are indicated by dashed lines. A plateau is observed, eventually followed by a decline phase. (B) Same simulation, but cumulative infection numbers are plotted. (C) Schematic illustration of a typical (uncut) spatial network. (D) Schematic illustration of the cut network, which results in the existence of one-dimensional infection corridors. (E) The cut network (red) superimposed onto the uncut network. (F) Same type of simulation as in part (A), but social distancing is initiated when different numbers of infected individuals are reached: 100 (as in part A), 200, 300, 400, 500, 500, 700. These are again averages over 900 simulations, and standard errors are indicated by dashed lines. (G) Same, but cumulative number of infections are plotted. Parameters were as follows. P_inf_=0.0001min^-1^ per edge; P_rec_=0.0001min^-1^; P_death_=0.00005min^-1^.

These plateau dynamics are explained as follows. When the network is relatively well-connected (pre-social distancing), the infection can spread in two dimensions (Figure 3C), resulting in power law growth, where the number of infections grows quadratically in time, see e.g. the classical results of [30, 31]. When the network connections are significantly cut, however, the remaining social pathways along which the infection can spread turn out to be significantly longer, resembling one-dimensional corridors (Figure 3D,E). Infection spread across a one-dimensional graph results in a constant number of new cases per day. Although the number of new cases remains roughly constant over time, however, the infection is still spreading through the community. Over time the infection spread reaches the end of these one-dimensional paths, at which point further spread cannot occur anymore and the infection levels start to decline. Therefore, the plateau phase can be explained by a transition from 2-dimensional to 1-dimensional infection spread, and indicate that the infection is now spreading towards a dead end.

Figure 3F shows a number of repeats of such simulations, but social distancing is implemented at different percentages of infected individuals, ranging from low to high. Interestingly, we observe that the plateau phase becomes less pronounced the more the infection has spread when social distancing is implemented. For the simulation where social distancing is implemented at the largest percentage of infected individuals, we observe a brief shoulder phase, followed by a relatively rapid decline of infection cases. This might explain why the plateau tends to be observed only in those locations that started social distancing early enough to prevent extensive infection spread.

While we have demonstrated the dynamics for a particular parameter combination and spatial network configuration, Figure S9 shows similar dynamics for different parameters, and also if a less severely cut network during social distancing is assumed.

Next, we investigated these dynamics in the other networks. The scale-free Barbasi-Albert network is much more interconnected without any spatial components. Hence, upon cutting connections, a transition to a roughly 1-dimensioal spread does not happen, and plateau dynamics are not observed (Figure S8). As expected, the hybrid network reproduces the plateau behavior, but to a lesser extent than the spatial network (Figure S8).

Last but not least, the network simulations indicate that immunity of recovered individuals is an essential component of the plateau behavior. This is illustrated in Figure S13, which compares the effect of social distancing on infection spread dynamics in simulations that do and do not assume that recovered individuals are immune, assuming the spatial network.

Without the assumption of immunity, the plateau behavior is not observed, and the number of infected individuals during the phase of social distancing reaches a significantly higher peak (Figure S13). Based on these findings, we hypothesize that the beneficial effect of social distancing is noticeably enhanced by immunity. The reason for this model behavior is that recovered, immune individuals, even if not very prevalent in the population, can provide local roadblocks for infection spread, which contributes to the infection paths being more one-dimensional rather than two-dimensional.

### Infection spread upon relaxation of social distancing

Economic considerations require an eventual relaxation of social distancing measures, which has by now commenced in Europe, the United States and elsewhere. It is thought that this will enable a resurgence of infection levels, which is also referred to as a second wave. We investigated what our network models predict in this regard. This is a topic that has previously been analyzed with ordinary differential equation SIR models [8]. This study sought optimal social distancing schedules, where the starting time as well as the extent and duration of social distancing was varied with the aim to find the schedule that minimized over the first and the second peak of infection levels. One finding was that a longer duration of social distancing lowered the peak of the second wave of infection spread following the relaxation of social distancing measures [8]. Similar behavior is also observed in our network models, but added insights can be obtained arising from the existence of the plateau phase when social distancing is implemented. In agreement with the previous work [8], the network models also indicate that a longer duration of social distancing leads to a lower second wave. It does so, however, in a stage-wise manner (Figure 4A,B). While the dynamics are in the plateau phase, a later return to the fully connected network does not significantly decrease the subsequent infection peak (Figure 4A) or the final epidemic size (total number of individuals infected since the beginning of the epidemic, Figure 4B). Once the dynamics have entered the post-plateau decline phase, however, both the infection peak upon return to the fully connected network, as well as the final epidemic size, are noticeably reduced (Figure 4A,B). The model thus gives rise to an important policy suggestion: If plateau-like dynamics are observed during social distancing, it pays off to wait for the transition to the decline phase before relaxing the non-pharmaceutical interventions, such that both future public health burden and economic hardship are reduced.

**Figure 4.**
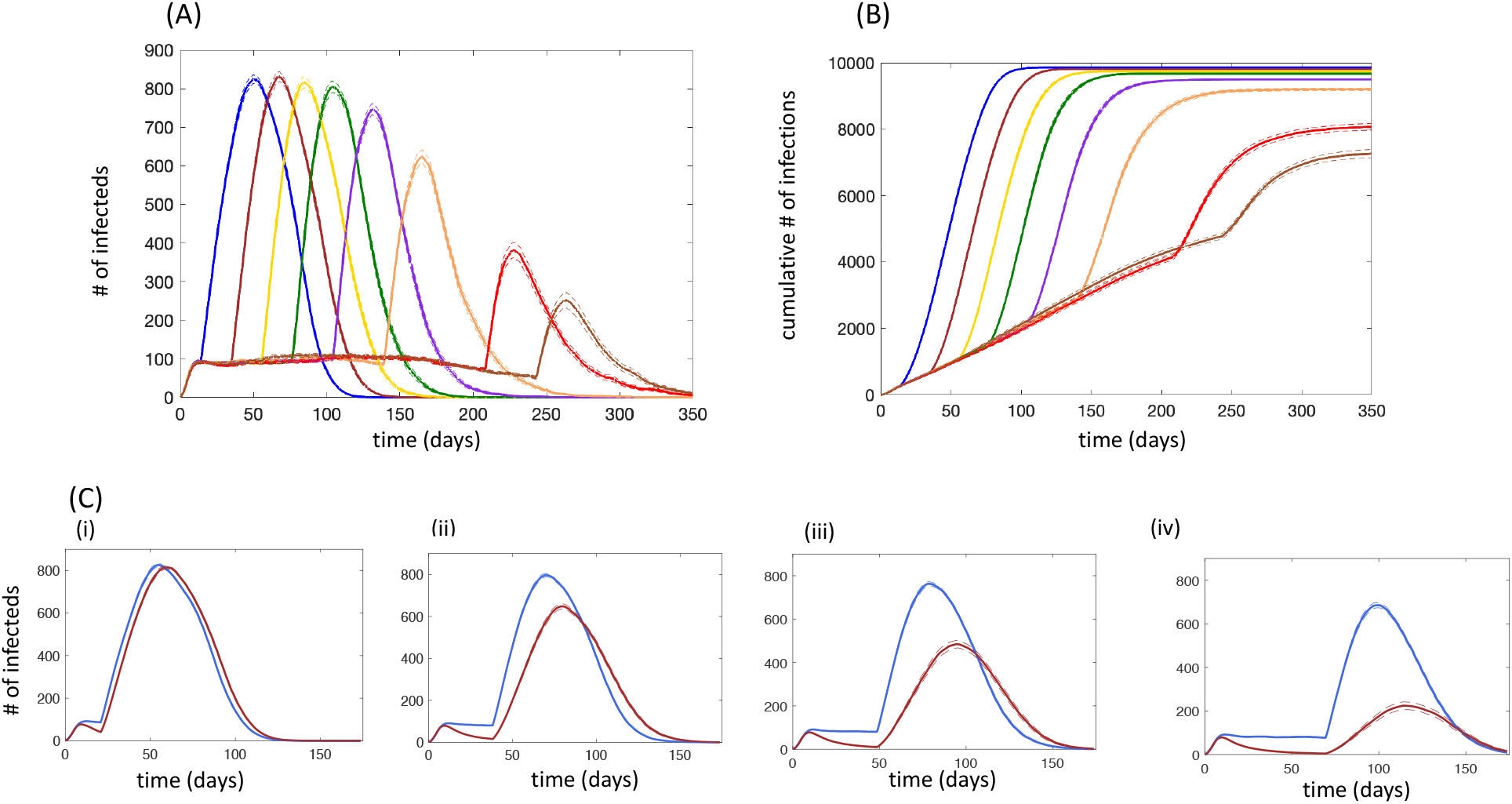
Infection spread dynamics when social distancing is relaxed in the spatial network model. (A) The number of infected individuals is plotted against time. Simulations start with the uncut network. When the infected population reaches size 100, ½ of randomly chosen edges are removed. At different times following the cut, the simulation reverts back to the original network. This results in a renewed wave of spread, and we let the infection spread in the simulation without further network cutting. Generally, a later return to the uncut network leads to a lower peak of the renewed growth. This reduction, however, is very minor, unless the return to the uncut network occurs when the infection levels are already in the decline phase during social distancing. The average over 900 simulations is shown. Standard errors are shown by dashed lines. (B) Same, but cumulative infections over time are shown. (C) Dynamics of the second wave after return to the uncut network, comparing different degrees of social distancing. The blue curve assumes that 50% of the connections are cut during social distancing. The orange curve assumes that 65% of the connections are cut during social distancing, i.e. distancing is stricter. Panels (i) – (iv) show return to the uncut network after longer durations of social distancing. Generally, stricter social distancing leads to a lower peak of the second wave of infections. For final epidemic size, see Figure S10. Each curve represents the average over 900 simulations. Standard errors are shown by dashed lines, which in some cases are too small to see. P_inf_=0.0001min^-1^ per edge; P_rec_=0.0001min^-1^; P_death_=0.00005min^-1^.

The reason that the second peak and the final epidemic size are reduced if social distancing is relaxed during the decline phase of the dynamics is that both measures depend on the population sizes when non-pharmaceutical interventions are reduced, and in particular on the number of infected individuals at this time. Fewer infected individuals lead to a lower peak and a lower final epidemic size, and this is only achieved once the number of infected cases starts to decline during the phase of social distancing. The longer the social distancing phase is maintained during this decline, the lower the predicted second peak and the final epidemic size.

We also investigated how the magnitude of the second peak depends on the degree of social distancing, expressed by the degree to which the original network was cut (more cut connections correspond to stricter social distancing). In the spatial network, we find that less strict social distancing results in a higher second peak (Figure 4C) and in a higher final epidemic size (Figure S10), because the number of infected individuals by the end of social distancing is higher if the degree of distancing is less strict.

It is interesting that in SIR models based on ODEs, the opposite is observed: less strict distancing (expressed by a higher rate of infection) results in a lower second peak [8]. The reason is the assumption of perfect mixing in ODE models: less strict distancing leaves fewer individuals uninfected (and hence susceptible), and under a perfect mixing assumption, this significantly slows down the rate of infection spread following the end of social distancing. In the spatial network model, in contrast, the total number of uninfected individuals is less important due to limited connections in this model, and the number of infected individuals when social distancing ends is the main driving factor. Since a higher degree of mixing occurs in the scale-free Barabasi-Albert network (compared to the spatial network), the magnitude of the second peak depends in the same way on the degree of distancing as in the ODEs (Figure S11). The hybrid network displays intermediate behavior (Figure S12), where the relationship between the degree of distancing and the second peak depends on the timing of relaxation. If relaxation occurs relatively early, less distancing results in a lower second wave, similar as observed in ODE models. If relaxation occurs later, however, the relationship between the degree of distancing and the predicted second wave is non-monotonic (Figure S12).

## Discussion and Conclusion

We have used network models to interpret the dynamics of COVID19 spread during and after social distancing measures. The network models can account for two observations that cannot be easily reproduced with ODE-based SIR models: They predict that the infection spreads according to a power law, and further predict the presence of a prolonged plateau phase following the start of non-pharmaceutical intervention measures, if those are implemented relatively early during the epidemic. According to the network models, the plateau occurs because during strict social distancing, infection spread follows nearly one-dimensional transmission corridors, compared to spread in two dimensions before distancing. Another interesting finding was that according to the model, the plateau phase naturally transitions into a decline phase without any further increase in non-pharmaceutical intervention measures. If these are the dynamics that are happening in our communities, then a significant benefit can be achieved if the end of social distancing occurs once the dynamics are already in the decline phase. A premature end of interventions can lead to a higher second wave and a larger final epidemic size.

The network models further indicate that the time at which the interventions are initiated plays an important role, and this is supported by data. The plateau is predicted to be observed if social distancing measures are implemented early (such as in West Coast states), and it is predicted to be much less pronounced if those interventions are started later (such as in New York). This insight might add to our understanding of the heterogeneity in responses to social distancing measures that are found when comparing different locations.

Another interesting observation concerned the role of immunity for the success of social distancing. In our computer simulations, the plateau is only observed if we assume that recovered individuals are immune. In the absence of this assumption, the plateau dynamics are not observed and infection levels during non-pharmaceutical intervention measures are predicted to be significantly higher. Data about the level of protection in recovered individuals are currently not available.

Our results further emphasize some important differences between ODE and network-based modeling. While the traditional SIR ODE modeling paradigm is capable of reproducing many features of network infection dynamics, some aspects are not captured by the simplified ODE framework. For example, the shape of the infection growth is not correctly predicted by the ODEs, which becomes especially apparent from the absence of the plateau dynamics during interventions in ODEs. Further, ODE models suggest that the lower the infectivity parameter during the intervention, the higher the second infection wave and the resulting final epidemic size. This result is a consequence of the complete mixing assumption and it is weakened or disappears under a network modeling approach.

It is important to note that model results depend on model assumptions and that uncertainties remain in this regard. While we think that network models are more realistic descriptions of infection spread during non-pharmaceutical interventions than ordinary differential equations that assume perfect mixing, uncertainty remains about the exact contact structure in our societies, which can also differ from location to location (e.g. comparing urban with rural areas). It appears, however, that our results depend on the notion that cutting network connections can transform virus spread in 2 dimensions towards spread paths that are more one-dimensional in nature. It might be possible to test this notion with more detailed data on human contacts during social distancing.

Another source of uncertainty comes from the data that we are interpreting. While a wealth of information exists about confirmed COVID190 case counts in the US and around the world, these counts depend on testing levels, making it hard to compare different locations. The observation that a plateau is observed could in principle also be explained by the limited availability of tests as true infection levels rise. This is unlikely to be the case, however, given that the percent of positive tests is typically not near saturation. If we assume that the observed plateau dynamics reflect a real plateau of infection levels during social distancing, we conclude that network models can account for these details of the dynamics in a more accurate way than models based on ordinary differential equations.

## Data Availability

All data referenced in this manuscript are available at:
https://covidtracking.com/

